# The awareness and practice of female genital mutilation in Lagos State, Nigeria

**DOI:** 10.1101/2024.09.01.24312909

**Authors:** Esther T. Joshua-Raji, Itunu O. Dave-Agboola

**Author notes:** Correspondence should be addressed to E.T.J.

## Abstract

**Background:** The extensive practice of female genital mutilation (FGM) is common in Nigerian societies, and it is generally done on infants as early as eight days after giving birth to them in early childhood, but it is usually done before marriage or before the first child is delivered. FGM has apparently caused pain to women, varying from infection, haemorrhage to difficulty in childbirth, even emotional distress.

**Objectives:** This study wherefore investigated the awareness and practice of female genital mutilation in Alimosho local government area, Lagos state, Nigeria to recognize the influence of the practice of FGM and suggest how to stop the danger of FGM in the society. A cross-sectional study was done to appraise the awareness and practices of female genital mutilation among the residents of Alimosho local government area of Lagos state.

**Methods:** A self-structured questionnaire was given to 301 males and females selected randomly, and the data were analysed using the Chi-square statistical tool.

**Result:** The result shows that while there is a significant level of knowledge and awareness regarding Female Genital Mutilation (FGM) in this area, there remains a minute section of the populace that are unaware of the danger associated with the practice of FGM.

**Conclusion:** It is highly commendable that women and men should be educated by the health personnel or people who are well educated on the danger FGM imposes on their health in order to stop the practice of FGM, and the government should enforce the penalties on the violators of FGM laws.

## Introduction

Female genital mutilation, often known as FGM, is defined by the World Health Organization (2023) as the partial or entire removal of the external female genital organs and causing harm to the female external genitalia for cultural or other non-therapeutic reasons. Female genital mutilation or female circumcision is a practice that dates back thousands of years, and this practice is commonly performed on young girls in Nigeria between infancy and puberty in Nigeria (UNICEF, 2022). According to Omigbodun (2020), in some countries and among some ethnic groups in Nigeria, women have their labia minora, labia majora, and clitoris surgically removed, and their vulvae are closed except for a little opening that allows them to urinate and menstruate. As documented by UNICEF (2022), female genital mutilation (FGM) has been practiced on about 200 million women across 30 countries, including Nigeria.

The practice of female genital mutilation (FGM) is wide-spread in Nigerian societies and it is typically carried out on infants as early as eight days after delivery; In other regions, it may not be done in childhood but it is usually done before marriage, or before the birth of the first child, like among the Ibo people who live in the southern eastern part of the country (Omigbodun, 2020). Unfortunately, the procedure is often carried out by traditional birth attendants and local circumcisers who lack medical expertise and make use of unsterilized equipment such as razor blades, scissors, and broken glass which introduces infections to the female reproductive system (UNFPA, 2022). Female genital mutilation is largely hinged on culture, beliefs, and values (Omigbodun, 2020).

Nigeria has the highest absolute number of cases of female genital mutilation (FGM) in the world, accounting for about 25% of the projected 115-130 million circumcised women globally (UNICEF, 2022). In Southwest Nigeria, the prevalence of female genital mutilation is 30%, according to UNICEF (2022), and Lagos is the most densely populated city in South-west Nigeria with a large number of women of reproductive age, according to the World Population Review (2023). Therefore, there is a need to investigate the knowledge and practices of female genital mutilation among women in Lagos, Nigeria, and proffer justifications for the elimination of female genital mutilation based on the adverse health effects.

Here, we appraised the knowledge and practices of female genital mutilation among the residents of Alimosho Local Government Area of Lagos state. By gaining insights into women’s perspectives in Alimosho community, it could offer insights into what drives the continual practices of FGM. Existing literature reveals a gap in the perception of Lagos women about FGM. Investigating the perception of women that resides in Alimosho Local government area will enlighten the understanding of the public and could inform ideas on how to stop the practice. This study endeavors to bridge these knowledge gaps to enable public health professionals and advocates utilizing the study’s findings to develop effective strategies for positive change, irrespective of personal beliefs regarding the detrimental effects of FGM on women.

## Results

### Influence of the level of education on the willingness to practice of FGM

We asked the people of Alimosho Local Government if their level of education could have an influence on their awareness of FGM. It was discovered that the people that have a high level of education have more knowledge of the implications of the practice of FGM. Moreover, there was a significant number of respondents at all levels of education who are unwilling to practice FGM when analyzed by chi-square (P=0.000). This suggests that people with some level of education are aware of the implications of FGM and would not practice the act.

When analyzed by chi-square, the data shows the Chi-square value of 298.183 with P-value of 0.000. This indicates that the educational status of the residents of Alimosho Local Government Area is a significant factor that contributes to the practice of female genital mutilation. The stated hypothesis is rejected because education was found to be a significant factor that contributes to the knowledge and practice of FGM in Alimosho. The higher, the level of education, the less support the respondents showed for FGM.

### Influence of the type of religion on the willingness to practice of FGM

We wondered if the awareness and practice of FGM in Alimosho Local Government can be influenced by their type of religion. It was discovered that the two most prominent religions in the community were biased towards not practicing FGM. However, a few subsets still indicated interest in the practice of FGM. The data sampled more people practicing the Christian religion. The result presented in figure 2 was used to test the hypothesis that the residents of Alimosho Local Government Area would not significantly practice FGM. The table shows the Chi-square value of 247.581 with P-value of 0.000. This indicates that the residents of Alimosho Local Government Area are significantly willing not to practice female genital mutilation. The stated hypothesis is rejected because majority of the respondents in Alimosho local government area of Lagos state stated that they would not circumcise their daughters.

**Figure 1:**
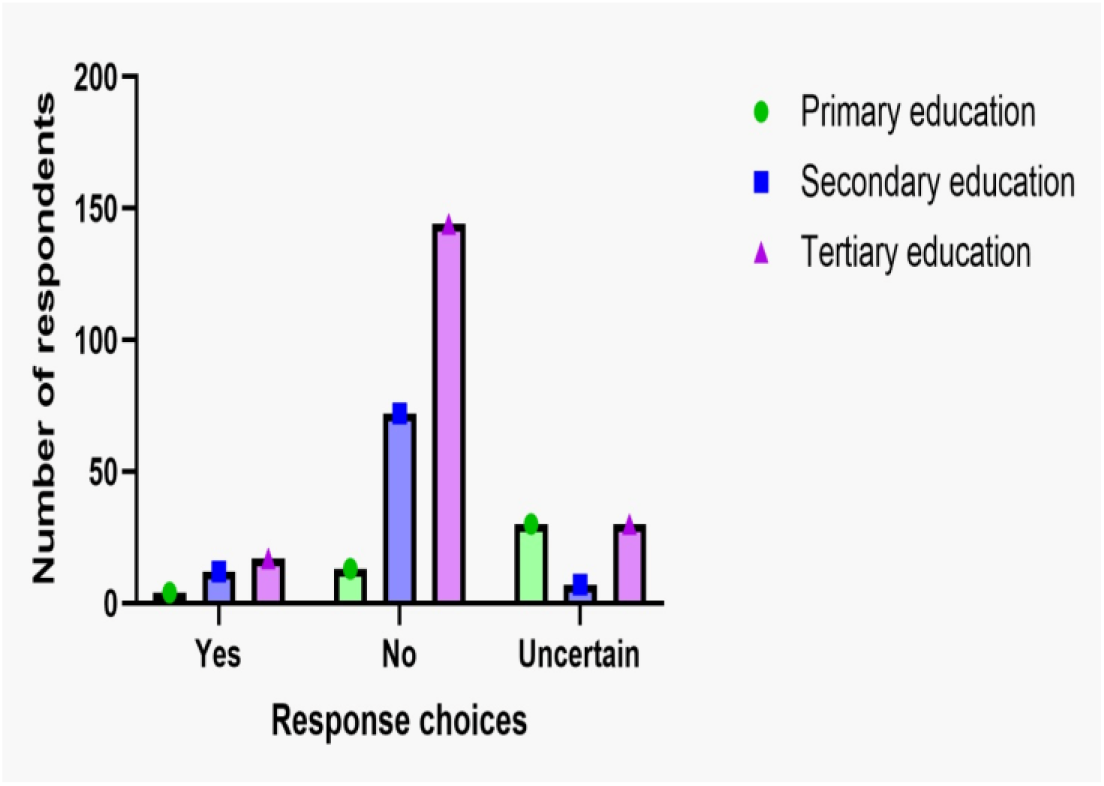
Level of education and willingness to practice. shows the number of respondents, and their response choices categorized based on their level of education. Data was analyzed by chi square to determine significance between groups.

**Figure 2:**
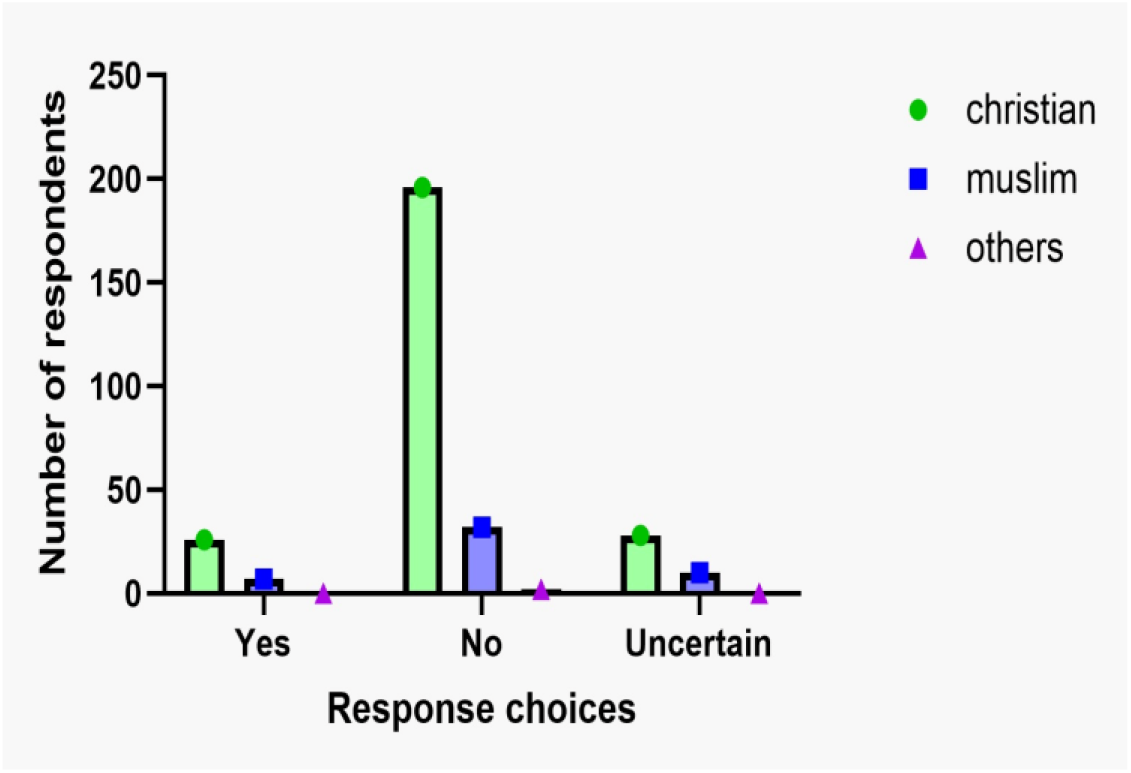
Type of religion and the practice of FGM. shows the number of respondents, and their response choices categorized based on their type of religion. Data was analyzed by chi square to determine significance between groups.

### Evaluating the willingness to practice FGM

We further investigated the overall willingness of the people of Alimosho Local Government to practice FGM. We identified significant avoidance of the willingness to practice FGM (P=0.000) as analyzed by Chi-Square. We also tested our hypothesis if cultural biases and personal belief can affect willingness. We found a very strong correlation between cultural practices and willingness to practice FGM. However, personal belief appears weakly correlated although it is significant (P=0.033).

The result presented in Figure 3 was used to test the hypothesis that the residents of Alimosho Local Government Area would not significantly practice FGM. The data analysis shows the Chi-square value of 247.581 with P-value of 0.000. This indicates that the residents of Alimosho Local Government Area are significantly unwilling to practice female genital mutilation. The stated hypothesis is rejected because the majority of the respondents in Alimosho local government area of Lagos state stated that they would not circumcise their daughters.

**Figure 3:**
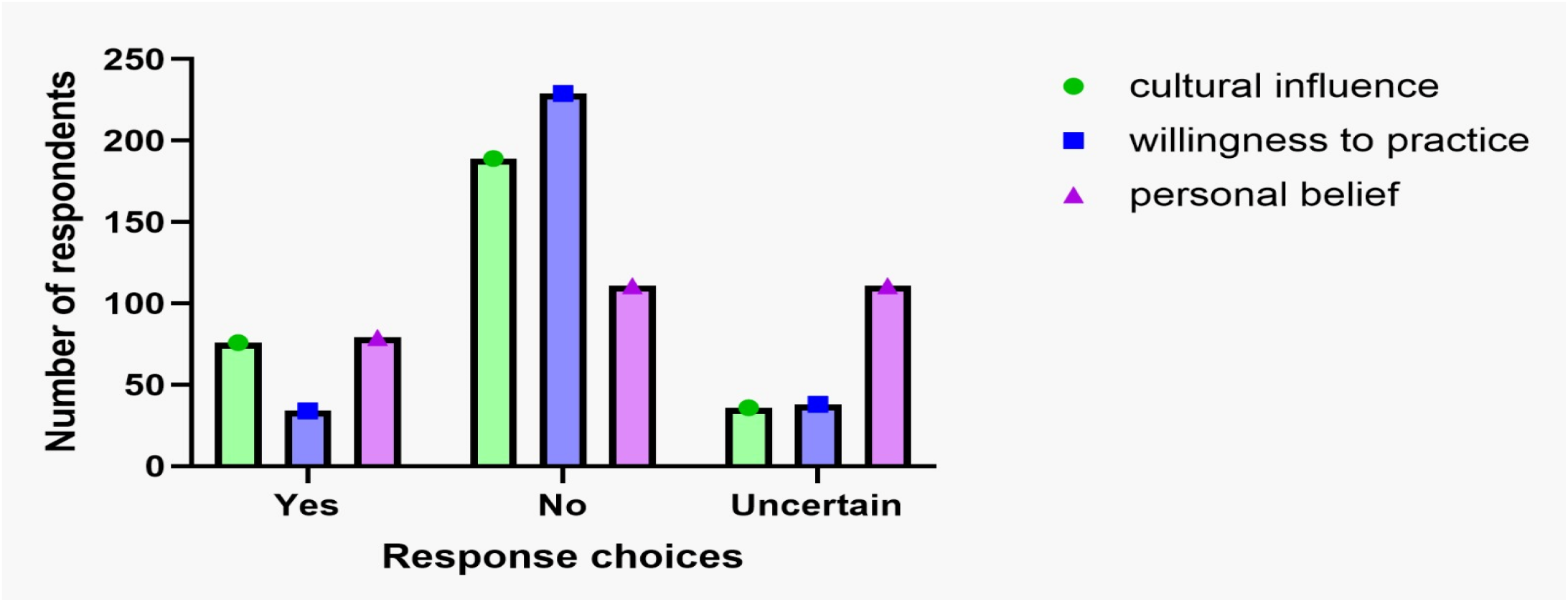
Type of religion and the practice of FGM. shows the number of respondents, and their response choices categorized based on their willingness to practice FGM. The effect of culture and personal belief on willingness to practice FGM was investigated. Data was analyzed by chi square to determine significance between groups.

Also, the result presented in Figure 3 was used to test the hypothesis that the personal belief of the residents of Alimosho Local Government Area is not a significant factor that contributes to the practice of FGM. The data shows the Chi-square value of 6.804 with P-value of 0.033. This indicates that the personal belief of the residents of Alimosho Local Government Area is a significant factor that contributes to the practice of female genital mutilation. The stated hypothesis is rejected because personal belief was found to be a significant factor that contributes to the knowledge and practice of FGM in Alimosho Local Government Area.

The final hypothesis tested was shown in figure 3. We asked if culture would not be a significant factor that contributes to the practice of FGM among the residents of Alimosho Local Government Area. The data shows the Chi-square value of 125.508 with P-value of 0.000. This indicates that culture is a significant factor that contributes to the practice of female genital mutilation among the residents of Alimosho Local Government Area. The stated hypothesis is rejected because culture is a significant factor that contributes to the practice of FGM in Alimosho.

## Discussion

This study shed light on the awareness and practice of female genital mutilation in Alimosho Local. An assessment of the level of knowledge of female genital mutilation among Alimosho Local Government residents revealed evidence of adequate knowledge, however, a minority of the residents remained ignorant. This observation is consistent with a report from an FGM campaign in Ebonyi State, Nigeria where a group of FGM proponents strongly upheld their beliefs that it is the only way to keep their wives from prostitution (Ikea, Onuh, and Emela, 2021). While majority of the residents of Alimosho Local Government stated that they would not circumcize their daughter, it is unsurprising that some would undoubtedly practice FGM. According to Koski & Heymann (2017) who reported a thirty-year trend analysis of FGM prevalence across 22 countries, they noted that the prevalence of FGM in certain countries is largely due to inadequate strict policies. The group of individuals that indicated the desire to circumcize their daughters were able to make this decision, for the most part, because of weak policies that govern the practice of FGM in Alimosho Local Government. These marginalized individuals lacking full knowledge of FGM may have been brainwashed by friends, neighbours, or religious leaders (Awolola & Ilupeju, 2019). Culture is a significant factor that contributes to the practice of female genital mutilation among the residents of Alimosho Local Government Area. This data corroborates the conclusions of a previous study, which posited that female genital mutilation (FGM) is entrenched within cultural and traditional contexts, with families assuming responsibility for determining whether their daughters undergo circumcision (Yount, et al., 2019). The existence of a minority that supports FGM is likely deeply rooted in culture, and it is usually passed down the family line.

This study also revealed that the educational status of the residents of Alimosho Local Government Area is a significant factor that contributes to the practice of female genital mutilation. Ameyaw et. al. (2020) showed that educational level correlates with knowledge and practice of FGM. The authors noted that the cohorts with higher education are fully aware of the consequences of FGM (Ameyaw et. al. 2020). The high level of knowledge about FGM from the respondents cannot be isolated from their educational level because the respondents who had a high knowledge of FGM were found to be mostly among the educated respondents. This corroborates the findings from another West African country (Seirra Leone) that educated people had more knowledge about FGM and its harmful effects and are therefore less likely to perpetrate FGM (Ameyaw, et al., 2020). This finding is also supported by another group in Ghana that postulates that educational status is directly proportional to the knowledge of FGM (Awuah, 2018).

The personal belief of the residents of Alimosho Local Government Area is a significant factor driving the practice of FGM. A significant proportion of the respondents belief that the decision about female genital mutilation (FGM) should be made inside the family unit. This group of residents upholds their personal belief above scholarly evidence and are unwilling to be educated about FGM. Consistent with the study carried out by Berg & Denison (2013), personal belief remained a critical factor perpetuating and hindering eradication of FGM. The outcome of the approach to FGM based on personal belief is in tandem with the result obtained from the religion of the respondent in the study. Awolola & Ilupeju (2019) highlighted in a scholarly review article that efforts should be geared towards using religious leaders to eradicate FGM. Personal belief can be shaped by religion, hence one’s religion may reflect one’s personal belief. Suffice to say that it is possible that the personal beliefs of the proponents of FGM may have been motivated by their religious practices.

## Summary

Female genital mutilation, commonly referred to as FGM, was defined by the World Health Organization as the partial or complete removal of the external female genital organs, causing harm to the female external genitalia for cultural or other non-therapeutic reasons. The practice of FGM was widespread in Nigerian societies, typically performed on infants as early as eight days after delivery. Nigeria had the highest absolute number of FGM cases in the world, accounting for about 25% of the projected 115– 130 million circumcised women globally.

In this study, we appraised the knowledge and practices of female genital mutilation among the residents of Alimosho Local Government Area of Lagos state. We investigated the knowledge and practices of female genital mutilation among the residents of Alimosho Local Government Area of Lagos state in specific areas, such as the health implications of female genital mutilation, factors that promote the practice of FGM, knowledge gaps of Alimosho residents on female genital mutilation. We hypothesized that the residents of Alimosho Local Government Area would have no significant level of knowledge of female genital mutilation, as well as certain factors such as culture, educational status, personal beliefs, and religion would not have a significant contribution to the practice of female genital mutilation.

Through an examination of the prevailing perception of women within the confines of Alimosho Local Government Area, the researcher has ascertained that while there exists a considerable degree of knowledge and awareness regarding Female Genital Mutilation (FGM) in this region, there still persists a segment of the population that remains uninformed about the inherent dangers associated with this practice. The study emphasizes the importance of considering diverse personal beliefs, cultural backgrounds, and religious perspectives when addressing the issue of female genital mutilation (FGM). The researcher has illuminated the personal awareness of women concerning FGM, thereby gaining insights into the underlying motivations behind the practice. Additionally, the study identified the factors that promote the practice of female genital mutilation in Alimosho Local Government Area of Lagos State and revealed the enduring motivations that had allowed FGM to persist for over 50 years despite being criminalized and subjected to various propaganda campaigns. The findings of this study held the potential to provide an understanding of how FGM may have held significant cultural value, contributing to its continual practice. Consequently, policymakers and interventionists could utilize this information to develop advocacy strategies that respect the community’s cultural context.

## Conclusion

From the findings of this study, it is concluded that the residents of Alimosho Local Government are largely informed and aware of the practice of Female Genital Mutilation (FGM), and the associated consequences. Nevertheless, a minority of the population residing in the Alimosho local government maintains the view that Female Genital Mutilation (FGM) is an imperative practice. It is noteworthy that the awareness and practice of FGM in Alimosho Local Government are much impacted by factors such as cultural and traditional norms, religion, and personal beliefs, as well as educational levels. Although personal beliefs have the weakest impact, compared to other factors, they are all considered critical factors influencing the practice of FGM. All the factors examined in this study shape awareness and practice of FGM. The knowledge of FGM appears to be widespread, but a minority that are strong proponents of FGM still exists, suggesting the practice of FGM in Alimosho Local Government is abated but not completely abolished.

## Data Availability

All data produced are available online through google form archive.

## Acknowledgements

We would like to express our gratitude to Dr. Dansu Tony, for his invaluable intellectual support, and all the participants that participated in this study. We thank Dr. Raji Joshua for data analysis.

## Materials and Methods

### Ethical Approval

Ethical clearance was obtained from the research ethics committee of the National Open University of Nigeria. The Local Government Area Authority of Alimosho also gave the study ethical approval and permission to enter communities for the purpose of data collection. Everyone who participated in the study received a detailed explanation of what the study entails, and they voluntarily signed an informed consent form. Those who declined to sign the consent forms were exempted from the research. The names of the research respondents were not written on the questionnaire and all the data collected was kept confidential to protect the privacy of the research respondents. Residents below 18 years of age were not included in this study to ensure that all the respondents are legally capable of providing informed consent.

### Data Collection

The sample of this study comprises 301 men and women of reproductive age living in the 6 administrative divisions of Alimosho Local Government Area of Lagos state. The samples were selected using a purposive sampling technique that targeted 50 participants from each administrative division in the Local Government. In the first stage, the purposive sampling technique was used to select Alimosho LGA in Lagos. This is because this LGA is densely populated and has a large number of women of reproductive age from diverse backgrounds, different levels of education, and different socioeconomic statuses. In the second stage of the sampling, the simple random sampling technique was used to select 50 respondents from each of the LCDA in Alimosho (Agbado/Oke-odo, Ayobo/Ipaja, Alimosho, Egbe/Idimu, Ikotun/Igando, and Mosan Okunola). In each of the LCDAs, women were randomly selected from house to house to be interviewed for this study with their consent. The data were collected by using a self-developed structured questionnaire.

### Data Analysis

Based on the quantitative methodology, descriptive statistics and a univariate analysis were carried out. The quantitative approach to social research was utilised to carry out studies on both of the aforementioned levels. To assess the socio-demographic factors, descriptive statistical analysis in form of percentages was conducted. A second level of analysis was performed with the use of univariate analysis to determine the relationship that exists between the socio-demographic features of the respondents and the independent variables (awareness and practice of female genital mutilation). It was vital to do this to evaluate the degree to which the socio-demographic features of the respondents predicted whether or not they are aware of the potentially detrimental effects of female genital mutilation. Inferences were thereafter made at a 0.05 level of significance.

